# Direct prediction of Homologous Recombination Deficiency from routine histology in ten different tumor types with attention-based Multiple Instance Learning: a development and validation study

**DOI:** 10.1101/2023.03.08.23286975

**Authors:** Chiara Maria Lavinia Loeffler, Omar S.M. El Nahhas, Hannah Sophie Muti, Tobias Seibel, Didem Cifci, Marko van Treeck, Marco Gustav, Zunamys I. Carrero, Nadine T. Gaisa, Kjong-Van Lehmann, Alexandra Leary, Pier Selenica, Jorge S. Reis-Filho, Nadina Ortiz Bruechle, Jakob Nikolas Kather

## Abstract

**Background:** Homologous Recombination Deficiency (HRD) is a pan-cancer predictive biomarker that identifies patients who benefit from therapy with PARP inhibitors (PARPi). However, testing for HRD is highly complex. Here, we investigated whether Deep Learning can predict HRD status solely based on routine Hematoxylin & Eosin (H&E) histology images in ten cancer types.

**Methods:** We developed a fully automated deep learning pipeline with attention-weighted multiple instance learning (attMIL) to predict HRD status from histology images. A combined genomic scar HRD score, which integrated loss of heterozygosity (LOH), telomeric allelic imbalance (TAI) and large-scale state transitions (LST) was calculated from whole genome sequencing data for n=4,565 patients from two independent cohorts. The primary statistical endpoint was the Area Under the Receiver Operating Characteristic curve (AUROC) for the prediction of genomic scar HRD with a clinically used cutoff value.

**Results:** We found that HRD status is predictable in tumors of the endometrium, pancreas and lung, reaching cross-validated AUROCs of 0.79, 0.58 and 0.66. Predictions generalized well to an external cohort with AUROCs of 0.93, 0.81 and 0.73 respectively. Additionally, an HRD classifier trained on breast cancer yielded an AUROC of 0.78 in internal validation and was able to predict HRD in endometrial, prostate and pancreatic cancer with AUROCs of 0.87, 0.84 and 0.67 indicating a shared HRD-like phenotype is across tumor entities.

**Conclusion:** In this study, we show that HRD is directly predictable from H&E slides using attMIL within and across ten different tumor types.

## Background

Homologous recombination repair (HRR) is a DNA repair mechanism that ensures genomic integrity after DNA double-strand breaks (DSB), which occur regularly during the cell cycle (1). Homologous recombination deficiency (HRD) results in defective DNA break repair, increased somatic copy number alterations and genomic instability, driving malignant transformation and causing cancer (2). Poly(ADP-Ribose)-polymerase (PARP) plays pivotal roles in base excision repair of single strand DNA breaks (SSDBs), which is a compensatory DNA repair mechanism in the context of HRD. In the setting of homologous recombination (HR) proficiency PARP inhibition results in the accumulation of unrepaired SSDBs. These can eventually convert to DSBs, which can be repaired via HR thus maintaining genomic integrity and cell viability. However in the case of a HRD tumor, PARP inhibition-induced DSBs are no longer repaired, resulting in direct cytotoxicity.. This phenomenon of *synthetic lethality* is the reason why HRD is an important biomarker to select patients for PARP inhibitor (PARPi) treatment in several tumor types, especially in breast, ovarian, prostate and pancreatic cancer (3–6). Prevalences of HRD varies according to the genomic definition of HRD and among tumor types, ranging from 0% in thymoma or thyroid cancer up to 70% in ovarian cancer(7). The use of PARPi has led to improved disease-free survival in multiple clinical trials by increasing platinum sensitivity in ovarian (OV) and breast cancer (BRCA), and other tumor types (8,9).

The success of PARPi therapy is mainly limited by the challenge of diagnosing HRD. Many different test strategies are available. The most robust test for HRD are oncogenic mutations in the Breast Cancer genes 1 and 2 (*BRCA1/2*) (10,11). However, this approach excludes patients without BRCA1/2-related deficiencies in the HR pathway (12). Moreover, other mechanisms such as epigenetic modifications, germline and somatic mutations of genes related or non related to the HRR pathway may cause HRD (13). Unfortunately, non-BRCA HR mutations have not been reliably shown to predict HRD or PARPi benefit in the clinic. Certain patterns of mutations, like the single base substitution 3 (SBS3) are also associated with a defective HR and therefore a potential biomarker (14,15). Finally, another strategy for detecting HRD is to look for the consequence of HRD rather than the cause. This approach uses whole genome sequencing single nucleotide polymorphism (SNP) array data to identify loss of heterozygosity (LOH), telomeric allelic imbalance (TAI) and large-scale state transitions (LST), also defined as a genomic instability score (GIS). This combined score has been validated in randomized clinical trials as predictive of PARPi benefit (16–18). Biologically, this methods provides a more comprehensive assessment of genomic instability caused by HRD, rather than scores exclusively based on mutation or HRR genes (Figure 1A). However, the GIS is not yet implemented in routine diagnostics in clinical workflows (11,12,19). Combining the different components of HRD using algorithms (e.g. scarHRD, HRDetect, CHORD) may be the gold standard to determine the genomic “scar” associated with HRD (20–22). A non-DNA-based way to determine HRD is using a functional test such as the RAD51 focus formation assays (23,24). U.S. Food and Drug Administration (FDA)-approved genetic tests for HRD typically rely on a combination of alterations in *BRCA1/2* genes and LOH (FoundationOne CDx, Foundation Medicine, Inc., Cambridge, MA) or GIS (myChoice CDx, Myriad Genetics Laboratories, Inc., Salt Lake City, UT) (10,11). However defining cut-off values for stratification between positive and negative cases is difficult (7,25). Taken together, the HRD testing landscape is highly complex. Many different tests coexist and they are not perfectly concordant. There is a high clinical need for a cheap, fast and standardized HRD test which captures a breadth of biological processes and not just alterations in individual genes. In this study, we hypothesized that the tumor phenotype as observed on histological whole slide images (WSI) of tumors reflects the HRD status and can be used to diagnose HRD.

**Figure 1:**
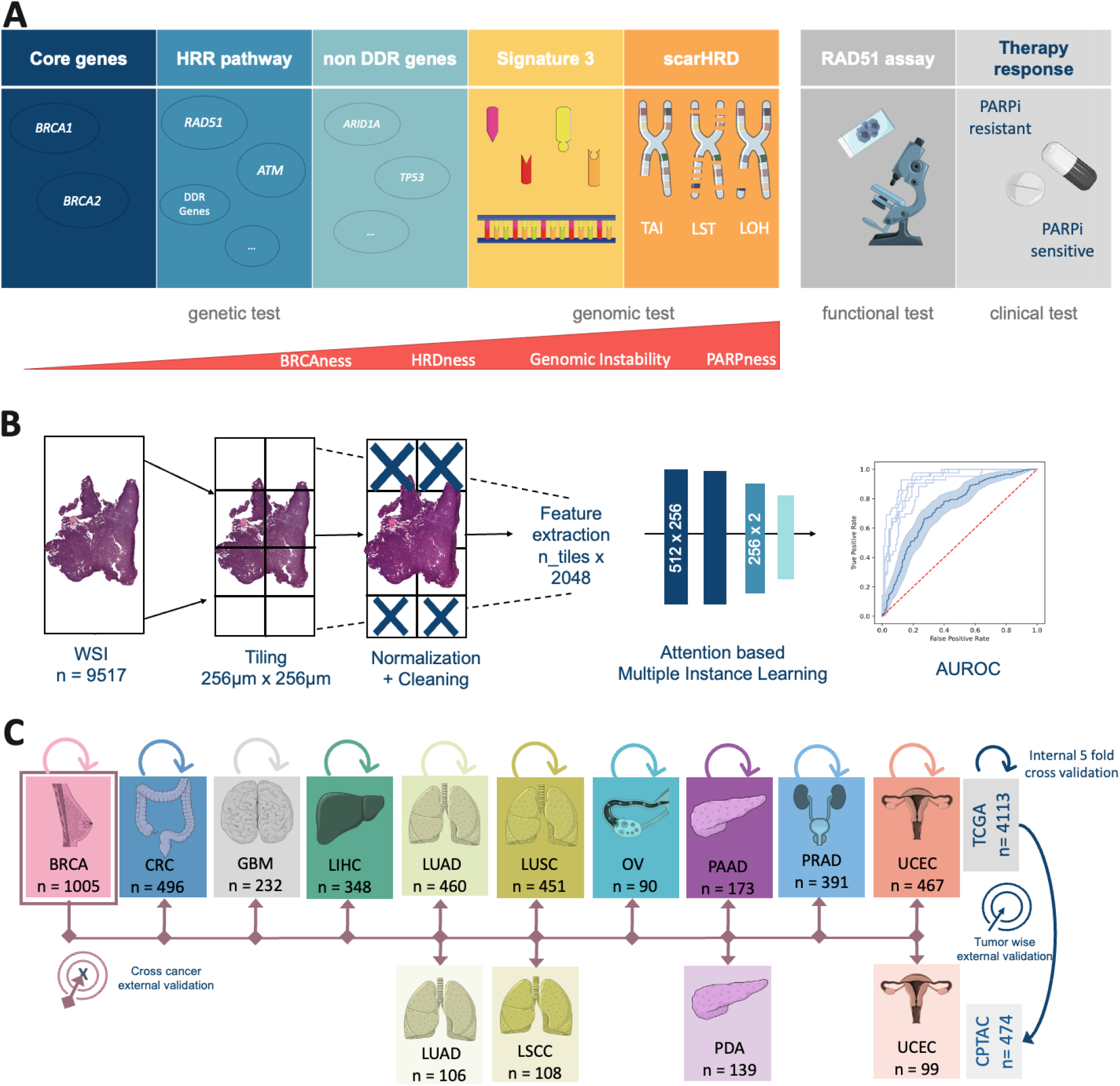
Experimental Design and Study overview. **(A)** Overview of the different Homologous Recombination Deficiency (HRD) scores, their content and assessment methods. **(B)** Workflow of our Deep Learning (DL) pipeline. A total of n=9517 Whole Slide Images (WSI) were processed and trained with an attention-based Multiple Instance Learning (attMIL) approach. The statistical endpoint was the Area under the receiving operating curve (AUROC). **(C)** Study design for the three main experiments (Internal 5-fold cross-validation, tumor-wise external validation and cross-cancer external validation) conducted and cohort overview for patients and tumor types included from The Cancer Genome Atlas (TCGA, n=4113 patients) and Clinical Proteomic Tumor Analysis Consortium (CPTAC, n=474 patients). Abbreviations: BRCA=breast cancer; CRC=colorectal cancer; GBM=glioblastoma; LIHC=liver cancer; LUAD=lung adenocarcinoma; LUSC/LSCC=lung squamous cell carcinoma; OV=ovarian cancer; PAAD/PDA=pancreatic adenocarcinoma; PRAD=prostate adenocarcinoma; UCEC=endometrial cancer; HRR=Homologous recombination repair. (This Figure was partly generated using Servier Medical Art, provided by Servier, licensed under a Creative Commons Attribution 3.0 unported license)

Deep Learning (DL) is an artificial intelligence (AI)-based technology which has emerged as a powerful method to quantitatively mine data from histological WSI of tumors in the last five years. DL enables us to detect genetic alterations directly from histopathological image data (26–28). Specifically, DL has been shown to detect single mutations(29,30), as well as phenotypic manifestation of DNA instability mechanisms such as microsatellite instability (MSI), just by processing scanned WSI of tumor tissue stained with H&E (31,32). Today, several DL systems to predict genetic alterations and clinical outcomes have received regulatory approval and are available for routine diagnostic use in Europe and the USA (33,34). Some smaller pilot studies have shown encouraging data for DL-based prediction of HRD from H&E WSI (35,36). However, HRD is a pan-cancer biomarker and DL has not been systematically used to diagnose HRD across tumor types directly from routine H&E pathology slides.

Therefore, in the present study, we developed a DL system to predict HRD status directly from H&E pathology slides. We used the state-of-the-art technology “attention-based Multiple Instance Learning” (attMIL) in a weakly supervised experimental setup, using no spatial labels or manual annotations whatsoever (28)ruth to train the DL system, we used the calculated scarHRD, one of the most comprehensive HRD scores which integrates a variety of genomic changes (Figure 1B). We trained and evaluated the DL classifiers by cross-validation in a large cohort of n=4,113 patients from The Cancer Genome Atlas (TCGA), comprising 10 types of solid tumors. The models were then externally validated on four cancer types in an independent validation dataset (n=474) in a tumor-wise and cross-cancer experimental approach (Figure 1C). Taken together, our experimental results provide direct evidence that HRD is detectable from routine histology in different types of cancer with DL.

## Methods

### Data Acquisition

In total data from 5,155 patients of 10 tumor types from The Cancer Genome Atlas (TCGA) and 573 patients from five tumor types from the Clinical Proteomic Tumor Analysis Consortium (CPTAC, Figure 1C) were obtained from https://www.cbioportal.org/. Accordingly, the cancer types included in the present study were breast invasive carcinoma (TCGA-BRCA n=1,058), colorectal cancer (TCGA-CRC n=580), glioblastoma (TCGA-GBM n=420, CPTAC-GBM n=99), liver hepatocellular carcinoma (TCGA-LIHC n=364), lung adenocarcinoma (TCGA-LUAD n=536, CPTAC-LUAD n=111), lung squamous cell carcinoma (TCGA-LUSC n=497; CPTAC-LSCC n=109), ovarian cancer (TCGA-OV n=520), pancreatic adenocarcinoma (TCGA-PAAD n=177; CPTAC-PDA n=153), prostate adenocarcinoma (TCGA-PRAD n=488) and endometrial carcinoma (TCGA-UCEC n=515, CPTAC-UCEC n=101, Supplementary Figure 1A,B). Image data and clinical data were available in TCGA-BRCA for n=1005, TCGA-CRC for n=496, TCGA-GBM for n=232, CPTAC-GBM for n=99, TCGA-LIHC for n=348, TCGA-LUAD for n=460, CPTAC-LUAD for n=106, TCGA-LUSC for n=451, CPTAC-LSCC for n=108, TCGA-OV for n=90, TCGA-PAAD for n=173, CPTAC-PDA for n=139, TCGA-PRAD for n=391, TCGA-UCEC for n=467 and CPTAC-UCEC for n=99, therefore leaving us in total with n=4,565 patients for the analysis (Figure 1C, Supplementary Figure 1A,B). Moreover, some figures were created using https://www.cbioportal.org/ (37,38). For additional experiments on *BRCA1/2* mutational status we retrieved data from Riaz et al previously published paper (39). Estrogen receptor data for the subgroup analysis was only available for n=661 patients in the TCGA-BRCA cohort.

### Image Preprocessing

WSIs were downloaded for the above mentioned cohorts from the GDC Portal (https://portal.gdc.cancer.gov/) and The Cancer Imaging Archive (TCIA, https://www.cancerimagingarchive.net/). Initially, the images were tessellated into patches with an edge length of 256 μm and a resolution of 224×224 pixels. Secondly, the patches for each cohort were color normalized using the Macenko spectral matching technique(40) to enforce a standardized color distribution across cohorts. To train the prediction models, we used our in-house open-source DL pipeline “marugoto” (https://github.com/KatherLab/marugoto) consisting of a self-supervised learning (SSL) model using a pre-trained ResNet50 architecture with ImageNet weights, fine-tuned pan-cancer on approximately 32.000 WSI to extract a 2048-dimensional feature vector for each patch per patient (41). To obtain patient-level predictions, 512×2048 feature matrices (MIL bags) are constructed by concatenating 512 feature vectors selected at random per patient and fed into an attMIL framework with the following architecture: (512×256), (256×2) with a subsequent attention mechanism (Figure 1B). (42,43)

### Calculation of HRD Scores

For the patient-wise calculation of HRD, single nucleotide polymorphism (SNP) data, generated by the Allele-Specific Copy number Analysis of Tumors (ASCAT) algorithm, was downloaded from the Genomic Data Commons (GDC) Portal: https://portal.gdc.cancer.gov/ (accessed 06/15/2022). In CPTAC, the respective data was only available for the CPTAC-3 cohort. The HRD score was calculated using the scarHRD (https://github.com/sztup/scarHRD), as described in previous studies (20,44). ScarHRD uses whole genome sequencing data in the form of SNP arrays to calculate the three subscores LOH, LST and TAI. The sum of these subscores makes up the patient-wise HRD score (Figure 1A). The cut-offs of the different subscores have been previously defined by Abkevich et al. for LOH, Popova et al. for LST and Birkbak et al. for TAI (16–18). Adding up the LOH, LST and TAI scores, patients can be divided into HRD high (HRD-H) and HRD low (HRD-L) at a cut-off of 42 (7). All patients in the CPTAC-GBM cohort were HRD-L. Hence, we excluded them from further analysis (Supplementary Figure 1A,B).

### Experimental Design

In our study, we performed three main experiments (Figure 1B). To assess the baseline predictability of HRD from routine histology, we first trained an HRD classifier in a within-cohort approach using five-fold-cross-validation within each of 10 tumor entities mentioned above in the TCGA cohorts (internal validation). This was achieved by randomly splitting each cohort on the level of patients, creating non-overlapping training and test sets for model training. The ratio for splitting the training and testing set was 80:20 of the entire dataset, and the training and validation set was split 75:25 of the training set. Thus, the absolute split for training, internal validation and internal testing was 60, 20 and 20, respectively. Five different models were trained until each part was used as a test set once. Thus, no data leakage from the test set to the training set occurred. This process was repeated individually for each cancer type in the TCGA cohorts. A weighted cross-entropy loss function was used to assist the model with the imbalanced dataset. Secondly, we deployed the five models trained in the first experiments on the same tumor type from the CPTAC cohorts as an external validation. By utilizing this approach, we circumvent any potential claims of selecting the model with the highest AUROC in the external validation. Lastly, we trained an HRD classifier on the TCGA-BRCA cohort, which had the highest number of patients, and deployed it on all other TCGA cohorts (CRC, GBM, LIHC, LUAD, LUSC, PRAD, PAAD, OV, UCEC) as well as on all CPTAC cohorts (LUAD, LSCC, PDA, UCEC). In our study, we aimed to evaluate the performance of the models using the AUROC, which is commonly used for assessing the accuracy of binary classification tasks. Our primary statistical endpoint was the AUROC +/-95%-confidence interval (CI) and Area under the precision recall curve (Supplementary Table 1). To further assess the performance of each model, we used a two-sided t-test to compare the patient-level prediction scores between the HRD-H and HRD-L patient groups as defined by the ground truth and report the p-values, assuming a significance level of 0.05 as statistically significant, without correction for multiple testing (Supplementary Table 1). As a final step to obtain a more profound understanding of the TCGA-BRCA cohort, we uploaded our custom HRD-H and HRD-L ground truth and predicted subgroups in cbioportal to examine the characteristics of these cases in the TCGA-BRCA PanCancer Atlas cohorts.

### Explainability

To visualize the output of our model, we created high resolution heat maps that show the spatial distribution of our model’s attention and prediction scores on the WSI. Therefore, using RetCCL convolutional neural network image feature vectors for 32×32 pixel fields were extracted from the WSI. We then calculated attention and classification scores for each image region and normalized them across the distribution of scores within each patient cohort. Based on these scores, color heatmaps for each patient, with red indicating high attention or a positive classification and blue indicating low attention or a negative classification were generated. To ensure interpretability of the underlying morphology together with the attention and classification scores, we separately reconstructed the final attention and classification heatmaps by blending the raw color heatmaps with the image features. This approach allows us to interpret the output of our model in a way that is easy to understand and provides insight into the underlying morphology of the tumor.

## Results

### HRD is predictable from histology with attMIL

First, we investigated whether DL could predict HRD status from H&E types within 10 different types of cancer from the TCGA cohort. We used cross-validation on the level of patients to train and test an attMIL-based DL model within each cohort. In our dataset, the prevalence of HRD ranged from 3% in glioblastoma (GBM) up to 63% in OV (Supplements Figure 1C). We found that in five out of 10 cancer types, the mean prediction AUROC was above 0.6, and the 95% CI of the fold-wise HRD prediction AUROCs remained above the null hypothesis of 0.5. Among these, HRD prediction reached statistical significance with a p-value below 0.05 in three cancer types: endometrial cancer (UCEC, AUROC 0.79+/-0.04, p=0.0008), breast cancer (BRCA, AUROC 0.78+/-0.02, p<0.0001) and lung adenocarcinoma(LUAD, AUROC 0.66+/-0.05, p=0.02; Figure 2A). AUPRC values are reported in the Supplementary Table 1. Prediction of HRD was not possible in LUSC, LIHC, GBM, as their prediction AUROCs did not exceed the baseline (0.55+/-0.04 0.56+/-0.14, 0.58+/-0.38) with CIs above the null hypothesis or p-values below 0.05 (Supplementary Figure 2 A-J, Supplementary Table 1). For the tumor types PAAD, OV and PRAD, the AUROCs ranged from 0.58+/-0.22 to 0.6+/-0.09 to 0.76+/-0.22. Together, these data demonstrate that DL can predict HRD status from histology images alone in several tumor types.

**Figure 2:**
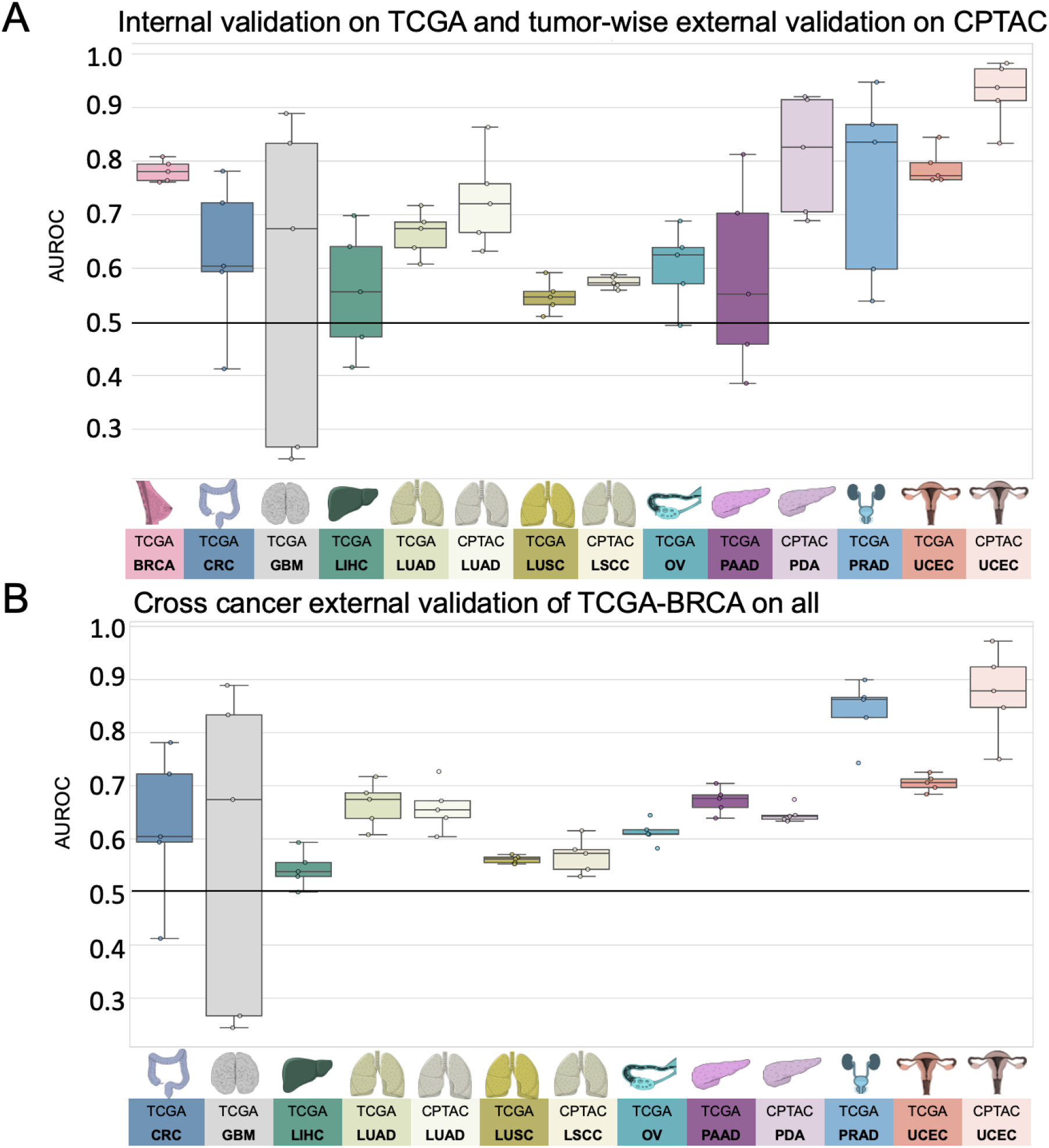
Comparison of Area under the receiving operating curve (AUROC) for internal and tumor wise external validation experiment models. Boxplot displaying the distribution for the AUROC for (A) internal 5-fold cross-validation experiment of The Cancer Genome Atlas (TCGA) and tumor-wise external validation on the Clinical Proteomic Tumor Analysis Consortium (CPTAC); (B) AUROCs for the cross-cancer external validation experiment of the TCGA breast cancer cohort (TCGA-BRCA) on the TCGA and CPTAC cohort. The horizontal line indicates the median, whereas each box represents the interquartile range (IQR) between the first and third quartiles. The whiskers extend from the box to the minimum and maximum values, considering 1.5 times the IQR. Abbreviations: BRCA=breast cancer; CRC=colorectal cancer; GBM=glioblastoma; LIHC=liver cancer; LUAD=lung adenocarcinoma; LUSC/LSCC=lung squamous cell carcinoma; OV=ovarian cancer; PAAD/PDA=pancreatic adenocarcinoma; PRAD=prostate adenocarcinoma; UCEC=endometrial cancer

### HRD is predictable from H&E histology with attMIL in an independent test set

A step that is germane to the successful development of DL models is external validation with WSIs from patient cohorts which are completely independent from the training set (45). Hence, for our external validation experiments, we deployed the classification models obtained from the cross-validation training on TCGA to analyze cohorts from the CPTAC dataset corresponding to the same cancer type. External validation cohorts in CPTAC were available for endometrial cancer (UCEC), pancreatic cancer (PDA), lung adenocarcinoma (LUAD), and lung squamous cell carcinoma (LSCC). In these external validation experiments, we noted that the prediction performance was highercompared to internal validation experiments. Once again, the best performance was obtained in UCEC, with an AUROC of 0.93+/-0.07, p=0.01. In LUAD, the performance increased in the external validation, yielding an AUROC of 0.73+/-0.11 and a significant p-value of 0.03. In the case of PAAD/PDA, where the internal validation was unsuccessful (internal validation AUROC 0.58+/-0.22), the external validation resulted in an improved AUROC reaching 0.81+/-0.14, albeit with a p-value of 0.07. Meanwhile, in LUSC/LSCC, no improvement in performance was observed in the external validation set compared to the internal training set (AUROC 0.57+/-0.01, p=0.23, Figure 2A, Supplementary Figure 2 K-N). Together, these data show that DL-based classifiers of HRD status generalize beyond the training cohort.

### A HRD classifier trained on BRCA detects HRD across various types of cancer

Lastly, we aimed to investigate if HRD-related morphological features in one cancer type can help to predict HRD status in another cancer type. This would point to a shared set of morphological features across cancer types, potentially allowing us to develop a pan-cancer pathology-based prediction system for HRD status. To test this, we applied our trained HRD classifiers in a cross-cancer experimental design. We used the breast cancer cohort TCGA-BRCA to train the HRD classification model because this cohort had the highest number of patients. Subsequently, we deployed this model on all other cohorts obtained from the TCGA and CPTAC datasets. Surprisingly, the BRCA-based model was able to significantly predict HRD from non-BRCA tissue in UCEC, PRAD and PAAD. For those three cohorts, the external deployment of a BRCA-based model resulted in higher prediction AUROCs than the respective internal validation experiments, reaching AUROCs of 0.70+/-0.02, p<0.001 in TCGA-UCEC, 0.84+/-0.07 and p=0.004 in TCGA-PRAD 0.67+/-0.03, p=0.2 in TCGA-PAAD, 0.87+/-0.1 p=0.05 in CPTAC-UCEC and 0.65+/-0.02 p=0.26 in CPTAC-PDA, respectively (Figure 2B). In the tumor types LUAD and OV, the AUROCs remained with 0.62+/-0.03 for TCGA-LUAD, 0.66+/-0.06 for CPTAC-LUAD and 0.61+/-0.03 in TCGA-OV in a similar range to the internal validation results (Supplementary Figure 3A-M). Together, these data show that a classifier trained on breast cancer can predict HRD status from histology in other tumor types, indicating a shared “HRD morphology” between tumor types.

### Molecular and histomorphological characterization of TCGA-BRCA HRD-H and HRD-L cases

Finally, we investigated which molecular and morphological patterns were associated with ground truth and DL-predicted HRD status. We used the TCGA-BRCA cohort to analyze this in detail, as this was the largest cohort. We observed that in the HRD-H subgroup, 45% were classified as basal-like breast cancers, 11% as HER2-enriched, 15% as Luminal A, and 26% as Luminal B. In contrast, only 7% of the cases in the HRD-L subgroup were basal-like, 7% were HER2-enriched, 64% were Luminal A, and 18% were Luminal B (Figure 3A) (46). Within our predicted groups, we observed a similar distribution among the BRCA subtypes (Figure 3B).

**Figure 3:**
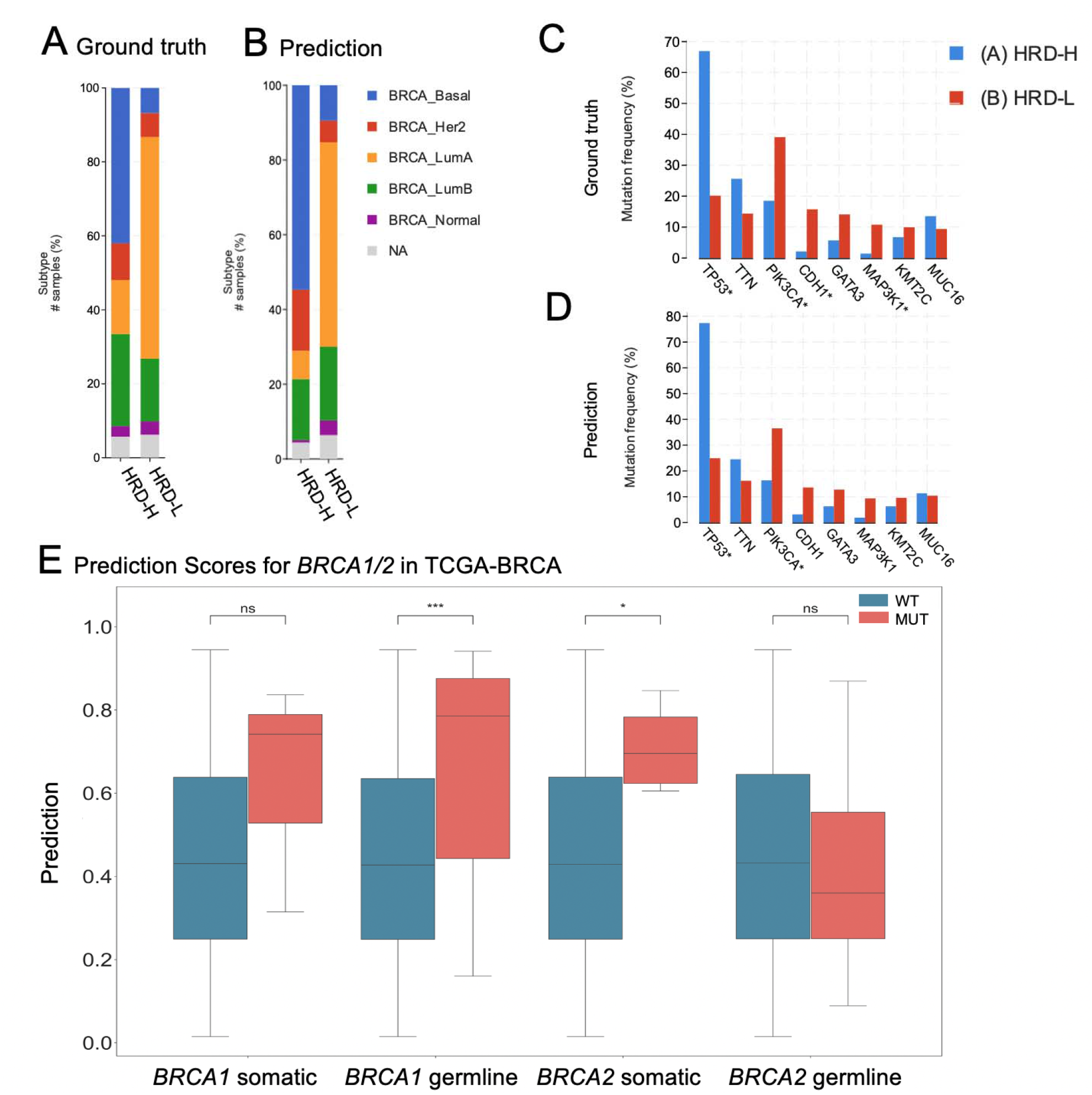
Molecular Characterization of The Cancer Genome Atlas breast cancer (TCGA-BRCA) cohort. (A) Distribution of breast cancer subtypes for the Homologous Recombination deficiency high (HRD-H) and low (HRD-L) ground truth subgroups. (B) Distribution of the breast cancer subtypes for the HRD-H and HRD-L Deep Learning (DL) predicted subgroups. (C) Alteration Frequency for several genes of the HRD-H and HRD-L ground truth subgroups. (D) Alteration Frequency for several genes of the HRD-H and HRD-L within cohort internal results prediction subgroups. (E) Grouped Boxplots comparing the Homologous Recombination Deficiency high (HRD-H) prediction scores with the mutational status (mutated=MUT, wildtype=WT) for the somatic and germline alterations of the *BRCA1/2* genes. The central line represents the median value, while the box ranges between the first and third quartile (IQR) and the whiskers extend to the lowest and highest values within 1.5 times the IQR. The y-axis represents the Deep Learning (DL) HRD-H prediction values. An independent t–test was performed to calculate the p-values: ns: p <= 1.00e+00 *: 1.00e-02 < p <= 5.00e-02 **: 1.00e-03 < p <= 1.00e-02 ***: 1.00e-04 < p <= 1.00e-03

To reassure that our model predicts HRD detached from phenotypic differences of estrogen receptor negative (ER-) vs. ER-positive (ER+) breast cancers we calculated the receiving operating curve (ROC) and precision recall curve (PRC) for the subgroups: ER+/*HER2*+, ER+/*HER2*-, ER-/*HER2*+, ER-/*HER2*-, indicating that HRD was also predictable with AUROCs of 0.66+/-0.3, 0.8+/-0.09, 0.72+/-0.43 and 0.62+/-0.11 (Supplementary Figure 4A-H). Our analysis of the mutational landscape of both HRD-H and HRD-L ground truth revealed that *TP53* had the highest alteration frequency with 67% in the HRD-H ground truth group, significantly higher than 20% in the HRD-L group, following alterations in the *TTN* (26% vs. 14%) gene. In contrast, the most enriched alterations in the HRD-L group were observed in the genes *PIK3CA* (39%) followed by *CDH1* (16%), *GATA3* (14%) and *MAP3K1* (11%), whereas the prevalences in the HRD-H group of *PIK3CA, CDH1, GATA3* and *MAP3K1* were 19%, 2%, 6% and 1%, respectively (Figure 3C). For the HRD-H prediction subgroup alteration frequencies for *TP53*, were significantly higher at 77% (Figure 3D). Such divergences were not as noticeable in the HRD-L prediction group. These findings suggest that there are notable differences in alteration frequencies between the two subgroups, which are consistent across both the ground truth and prediction data. Moreover, we compared the HRD-H prediction score to the alteration status of somatic and germline mutations in the *BRCA1/2* genes, whereupon we saw that there was a significant difference between the mutant and wild-type cases for *BRCA1* germline and *BRCA2* somatic mutations (Figure 3E). Methylation data indicated that the HRD-H group had most of its methylation alterations in the N-shore portion of the *BRCA1* promoter region, whereas those in the HRD-L group were mainly located in the S-shore portion (Supplementary Figure 4I). Lastly, we proceeded to investigate the histomorphological patterns associated with the presence of HRD through whole slide prediction heatmaps in CPTAC-UCEC (Figure 4A-C). Our findings revealed that high grade, fibrosis, hemorrhage and lymphocytic infiltration are consistent features predictive of HRD across various tumor types, as shown in Figure 4 for BRCA and UCEC, particularly in the top predicted HRD-H tiles for the top three patients. Fibrosis was observed in HRD-positive cases, particularly in BRCA (Figure 4D). Moreover, hemorrhagic necrosis especially adjacent to tumor tissue and tumor stroma was consistently observed as highly predictive areas in the true HRD-H cases across various cancer types. (Supplementary Figure 5, 6). In summary, these data show that known HRD morphology characteristics were found in our DL based top predicted HRD-H patients.

**Figure 4:**
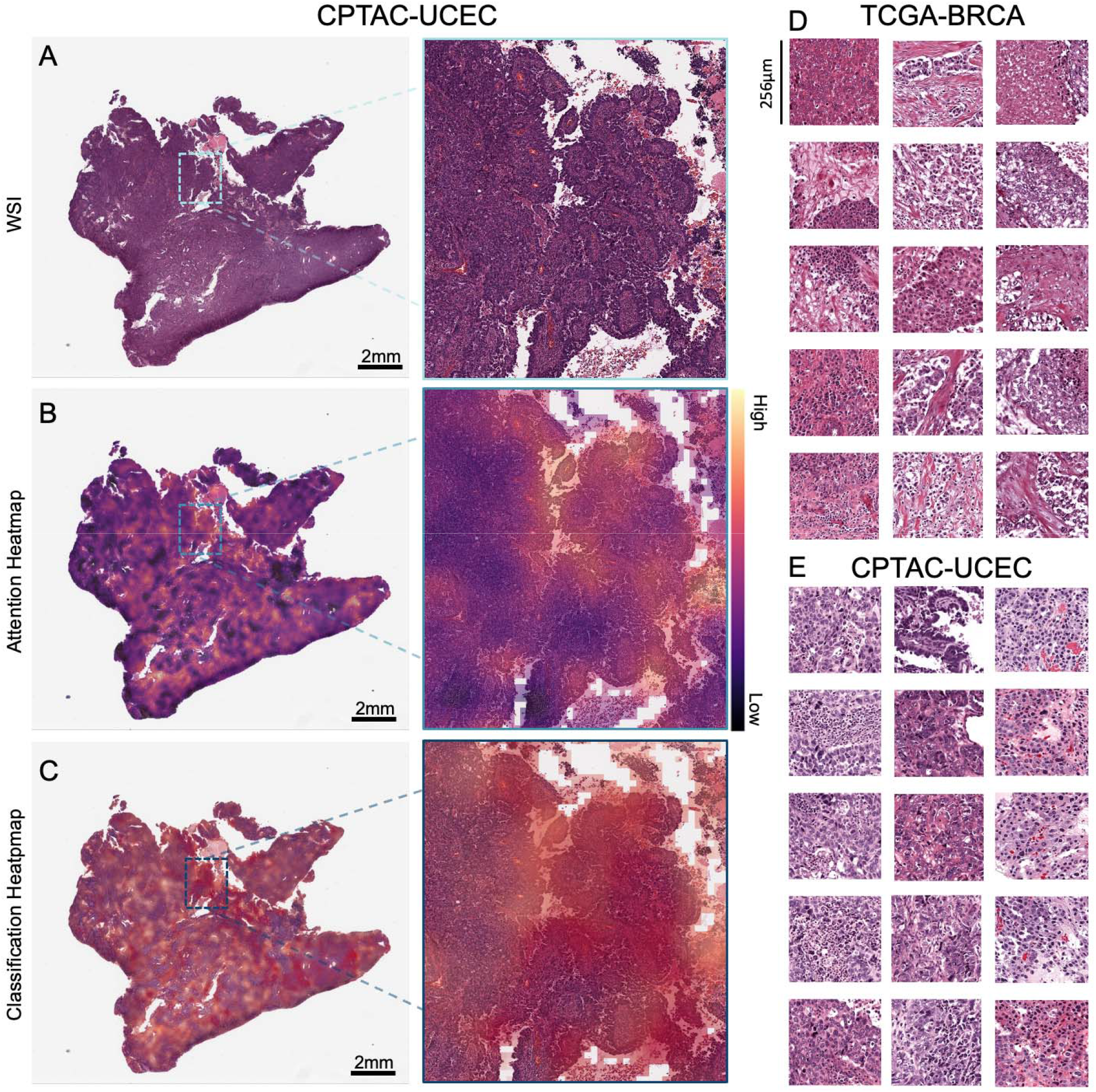
Visualization of predicted Homologous Recombination Deficiency high (HRD-H) tumor samples. (A) Whole slide image (WSI) of an HRD-H predicted patient (ID: C3L-00358-21) from the <u>Clinical Proteomic Tumor Analysis Consortium</u> (CPTAC) endometrial cancer (UCEC) cohort with magnification. (B) Attention heatmap for the same patient with magnification. (C) Classification Heatmap for the same patient with magnification. (D) Top predicted tiles for top three homologous recombination deficiency high (HRD-H) patients in The Cancer Genome Atlas (TCGA) breast cancer (BRCA). (E) Top predicted tiles for three HRD-H patients in the CPTAC-UCEC cohort.

## Discussion

HRD has recently emerged as an important pan-cancer biomarker for targeted treatment in solid tumors (11,47). The assessment of HRD clinically, albeit indicated for all patients with gynecological tumors, remains challenging. This is due to the given availability of different methods with limited agreement and whose logistic complexities and inherent costs pose significant hurdles for their adoption. In this light, a pan-cancer test of HRD by DL-based image analysis on pathology slides could be a useful pre-screening tool and reduce the load of genetic tests.

In this study, we demonstrated that DL can predict HRD status from H&E histology in different tumor types in both within-cohort and external validation experiments. Surprisingly, our findings revealed that a BRCA-based classifier could detect HRD from H&E slides across different tumor entities. As expected, the HRD prediction was significantly lower in tumors with a low prevalence of HRD. Our classifier has identified histomorphological characteristics such as hemorrhagic necrosis at tumor margins, lymphocyte infiltration, fibrosis, and high tumor cell density that are associated with HRD in BRCA(36). These findings validate the efficacy of our classifier. Moreover, despite having trained our classifier solely on BRCA, its consistent identification of HRD-associated morphological patterns across different tumor entities reiterates the value of our tool for broader applications. Compared to previous studies, we here show a pan-cancer DL-based prediction of a more comprehensive HRD score calculated from LOH, TAI, and LST as ground truth directly from H&E tumor slides. (35,36)

Our morphological analysis showed that UCEC or PAAD, achieved better predictive results compared to LUSC or LIHC, a trend previously observed in pan-cancer studies (30,48). In general, tumors with a complex structure, such as adenocarcinomas are morphologically susceptible to genetic alterations than solid tumors growing in rather syncytial patterns. HRD-positive tumors barely resemble glandular tissue anymore, which might be their main distinctive feature and therefore a potential explanation for this constellation. Additional studies with larger patient cohorts would be required to confirm this. A closer look at the TCGA-BRCA subgroups revealed that predicted HRD-H is more common in triple-negative breast cancer, which is known for its poor prognosis and resistance to conventional chemotherapy. In line with their ground truths, the majority of those patients were predicted to be HRD-positive by our classifier (Figure 3A,B) (46). Furthermore, clear molecular pathological differences were found in the two subgroups. Specifically, the HRD-H subgroup is characterized by *TP53* alterations, while the HRD-L subgroup has a higher frequency of PIK3CA alterations, suggesting an interactive effect between the *TP53* mutated cases and HRD-H patients (49,50). This is particularly true for *BRCA1* mutated cancers, where HRD-H was predicted significantly better than in *BRCA1* wildtype cases (Figure 3E) (51).

Recently, the EMA and FDA granted the first approval to use PARPi therapy for HRD positive ovarian cancer patients. Clinical trials with promising interim data are also underway for other tumor entities and further approvals are expected in the future. Despite the evident link between HRD and *BRCA1/2* mutations, it is now well established that the total number of HRD-positive patients significantly exceeds the total number of *BRCA*-mutated patients in various cancer types (22,52). The patients who fall into this diagnostic gap can be identified with comprehensive HRD testing, as proposed in our study.^41^ HRD testing would thus complement *BRCA1/2* testing as a biomarker test for PARPi use, such as with AI-based screening methods as applied here. Moving diagnostic routines towards phenotype-based instead of inconsistent, alteration-based HRD detection methods might extend our ability to identify patients who may benefit from PARPi and enroll them in clinical trials. Our study provides a proof of concept that there is indeed a pan-cancer preserved HRD morphology in histology slides which could potentially serve as an HRD marker. Prospective trials are needed to evaluate an AI-based HRD score as a biomarker to guide treatment decisions, potentially in a two-step approach leading to lower sequencing requirements and cost reduction.

## Limitations

Our study has several limitations. Firstly, the sample sizes of our cohorts, particularly the CPTAC dataset, are relatively small. Moreover, the variation within the distribution of HRD prevalences between tumor types can result in class imbalances. Although the effect of imbalanced datasets on the accuracy of our classifiers was addressed via weighing techniques during the model training process, this could still have an effect on the statistical power of the results, as well as the generalisability of our models to a larger population. We observed higher AUROCs in the external validation cohort, which may be attributed to the smaller size and higher class imbalance in the test set. Further studies with larger patient cohorts are required to validate our findings. Furthermore, the quality of the data from the TCGA and CPTAC cohorts may vary, thus potentially impacting the accuracy of our predictions.

## Conclusion

Our findings provide evidence that DL has the potential to not only contribute but improve diagnostic HRD testing, potentially saving time and costs as well as improving outcomes for patients by identifying subgroups who may benefit from targeted therapy. Current clinical practices face challenging factors such as high cost, time consumption, lack of availability, and inconsistency in HRD status screening methods. These logistic, analytic and financial challenges contribute to the partial identification of cancer patients who may benefit from PARPi therapy and to the limited genetic testing, which is further compounded by the panoply of HRD status assessment methods whose inter-assay concordance is limited. With the aid of AI, we have the opportunity to identify these subgroups and improve patient outcomes.

## Data Availability

The WSI, molecular and clinical data for TCGA and CPTAC cohorts are publicly accessible at https://portal.gdc.cancer.gov/ and https://www.cbioportal.org/ (accessed, 08 March 2022). Script for calculating the HRD score is available under https://github.com/sztup/scarHRD (accessed 06 June 2022). All other source codes can be downloaded under https://github.com/KatherLab/marugoto. Our calculated HRD score is publicly available in Supplementary Table 2. Moreover, our custom TCGA-BRCA HRD-H and HRD-L group can be accessed for the PanCancer Atlas cohort at https://www.cbioportal.org/ (Supplementary 3).

https://www.cbioportal.org/

https://github.com/sztup/scarHRD

https://github.com/KatherLab/marugoto.

https://www.cancerimagingarchive.net/

## List of Abbreviations

AI: artificial intelligence
ASCAT: Allele-Specific Copy number Analysis of Tumors
attMIL: attention-weighted multiple instance learning
AUROC: Area Under the Receiver Operating Characteristic curve
BRCA: breast invasive carcinoma
BRCA1/2: Breast Cancer genes 1 and 2
CI: confidence interval
CIOMS: Council for International Organizations of Medical Sciences
CPTAC: Clinical Proteomic Tumor Analysis Consortium
CRC: colorectal cancer
DL: Deep Learning
DSB: DNA double-strand breaks
ER-: estrogen receptor negative
ER+: estrogen receptor positive
FDA: U.S. Food and Drug Administration
GBM: glioblastoma
GDC: Genomic Data Commons
GIS: genomic instability score
H&E: Hematoxylin & Eosin
HR: Homologous recombination
HRD-H: HRD high
HRD-L: HRD low
HRD: Homologous Recombination Deficiency
HRR: Homologous recombination repair
LIHC: liver hepatocellular carcinoma
LOH: loss of heterozygosity
LSCC: squamous cell carcinoma of the lung
LST: large-scale state transitions
LUAD: adenocarcinoma of the lung
LUSC: squamous cell carcinoma of the lung
OV: ovarian cancer (OV)
PAAD: pancreatic adenocarcinoma
PDA: pancreatic adenocarcinoma
PARP: Poly(ADP-Ribose)-polymerase
PARPi: Poly(ADP-Ribose)-polymerase inhibitor
PRAD: prostate adenocarcinoma
PRC: precision recall curve
ROC: receiving operating curve
SBS3: single base substitution 3
SNP: single nucleotide polymorphism
SSDBs: single strand DNA breaks
SSL: self-supervised learning
TAI: telomeric allelic imbalance
TCGA: The Cancer Genome Atlas
TRIPOD: Transparent reporting of a multivariable prediction model for individual prognosis or diagnosis
UCEC: endometrial carcinoma
WSI: whole slide images

## Declarations

### Ethics statement

The experiments in this study were carried out according to the Declaration of Helsinki and the International Ethical Guidelines for Biomedical Research Involving Human Subjects by the Council for International Organizations of Medical Sciences (CIOMS). The present study also adheres to the “Transparent reporting of a multivariable prediction model for individual prognosis or diagnosis” (TRIPOD) statement.20. The Ethics Board at the Medical Faculty of Technical University Dresden (BO-EK-444102022) approved of the overall analysis in this study. The patient sample collection in each cohort was separately approved by the respective institutional ethics board.

### Data and Code availability

The WSI, molecular and clinical data for TCGA and CPTAC cohorts are publicly accessible at https://portal.gdc.cancer.gov/ and https://www.cbioportal.org/ (accessed, 08 March 2022). Script for calculating the HRD score is available under https://github.com/sztup/scarHRD (accessed 06 June 2022). All other source codes can be downloaded under https://github.com/KatherLab/marugoto. Our calculated HRD score is publicly available in Supplementary Table 2. Moreover, our custom TCGA-BRCA HRD-H and HRD-L group can be accessed for the PanCancer Atlas cohort at https://www.cbioportal.org/ (Supplementary Table 3).

### Author Contribution

CMLL, NOB, HSM, and JNK conceptualized the study. TS, CMLL, HSM and NOB curated the source data. MVT developed the source codes for the analysis. OSMEN, MG and CMLL conducted the experiments. CMLL interpreted the data and wrote the first draft of the manuscript. All authors revised the manuscript draft, contributed to the interpretation of the data and agreed to the submission of this paper.

## Supplementary Figures and Tables

**Supplementary Figure 1:**
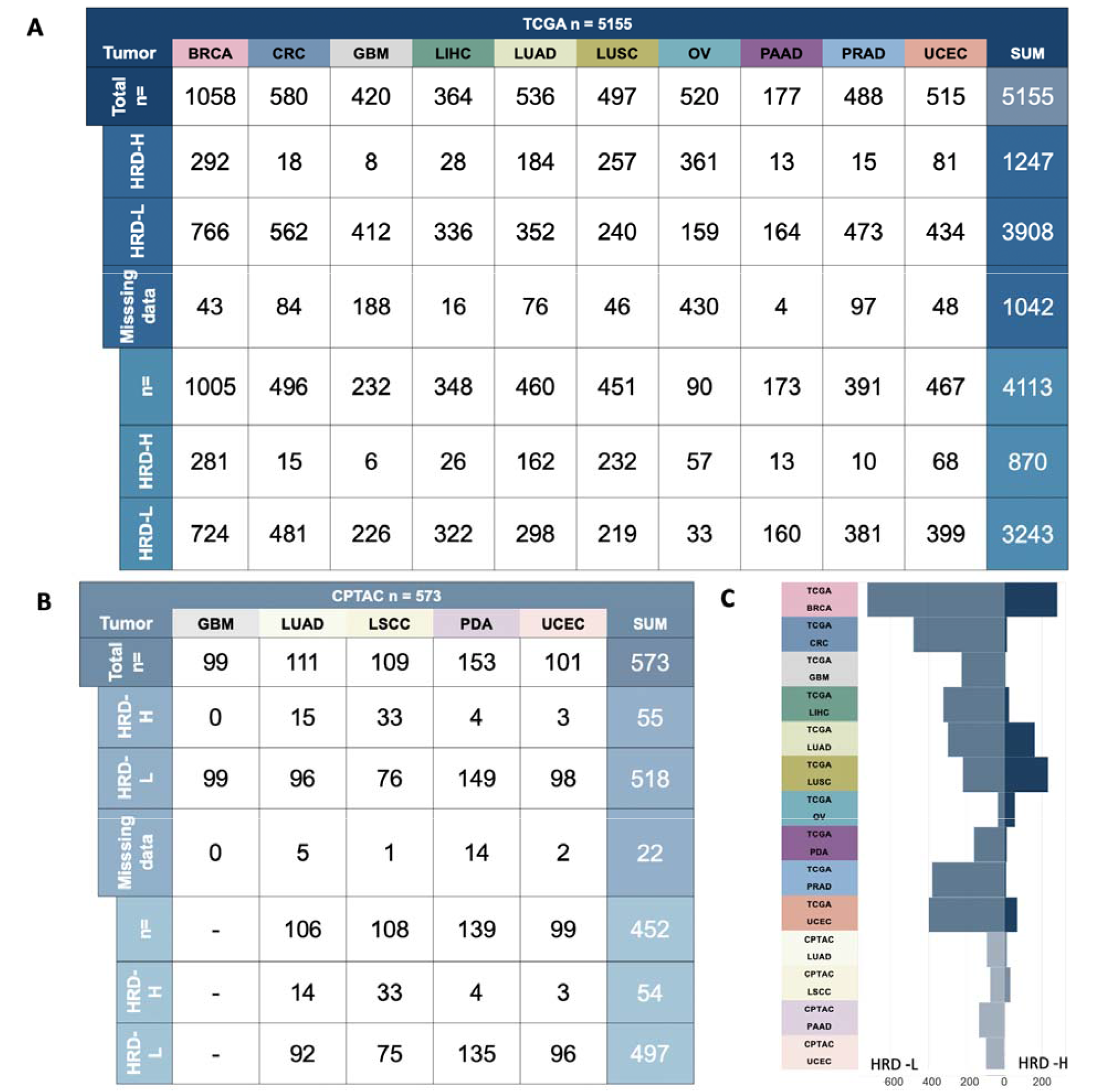
Homologous recombination deficiency prevalences across the cohorts. (A) Overview of the total patient count (n=573) in the CPTAC cohort before merging the image data with the molecular data and afterward. (B) Overview of the total patient count (n=5,155) in the TCGA cohort before merging the image data with the molecular data and afterward. (C) Distribution of the homologous recombination deficiency high (HRD-H) and low (HRD-L) patient number among the different tumor types of The Cancer Genome Atlas (TCGA) and Clinical Proteomic Tumor Analysis Consortium (CPTAC). Abbreviations: BRCA=breast invasive carcinoma; CRC=colorectal cancer; GBM=glioblastoma; LIHC=liver cancer; LUAD=lung adenocarcinoma; LUSC/LSCC=lung squamous cell carcinoma; OV=ovarian cancer; PAAD/PDA=pancreatic adenocarcinoma; PRAD=prostate adenocarcinoma; UCEC=endometrial cancer

**Supplementary Figure 2:**
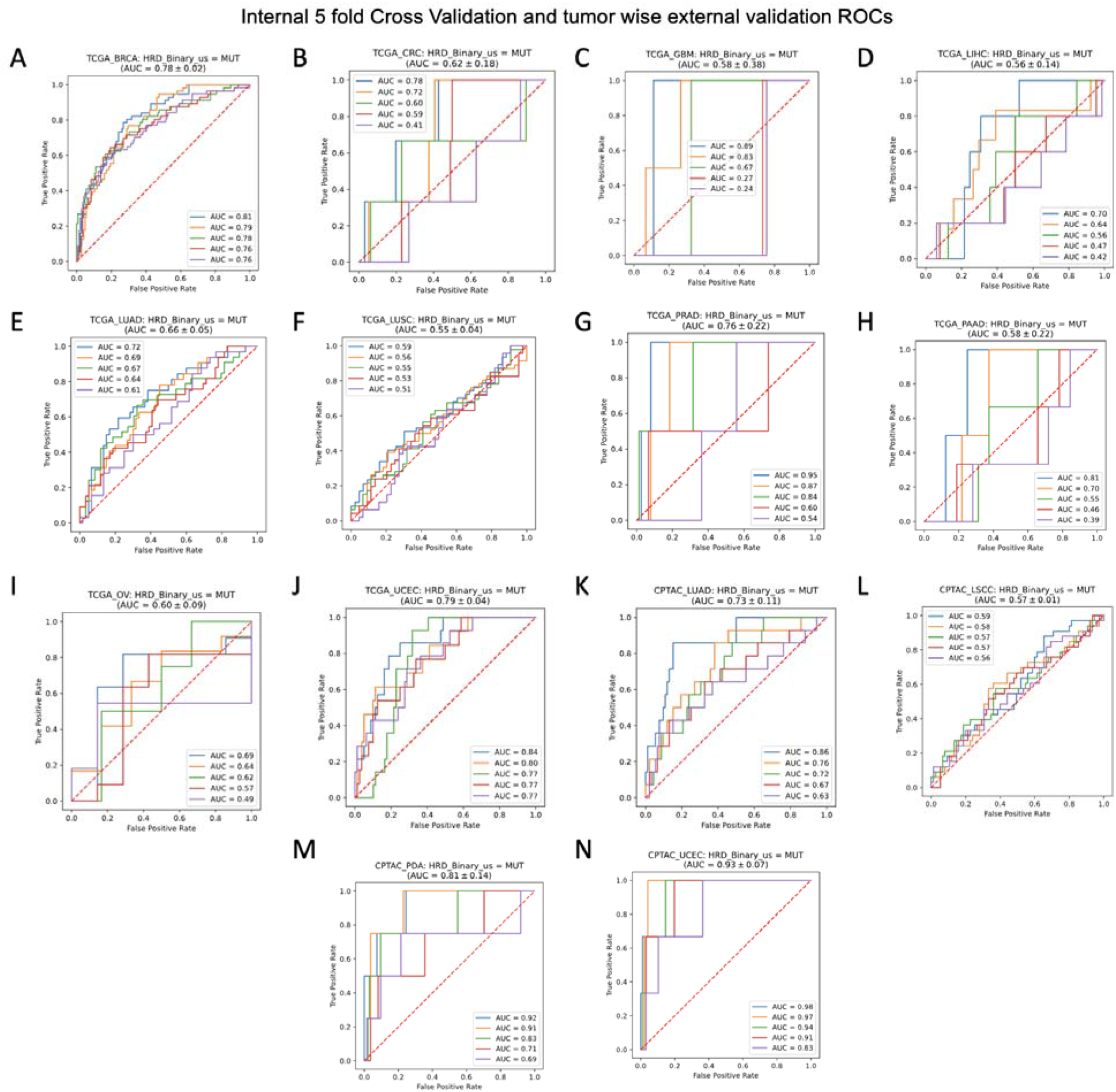
Receiving operating curve for the Internal Validation and tumor wise external validation. The Receiving operating curve (ROC) is shown for the five-fold internal cross-validation experiment for each of the models in The Cancer Genome Atlas (TCGA) for the Homologous recombination deficiency (HRD) binary score for (A) TCGA-BRCA, (B) TCGA-CRC, (C) TCGA-GBM, (D) TCGA-LIHC, (E) TCGA-LUAD, (F) TCGA-LUSC, (G) TCGA-PAAD, (H) TCGA-PRAD, (I) TCGA-OV, (J) TCGA-UCEC; Roc curves for the external validation on the Clinical Proteomic Tumor Analysis Consortium (CPTAC) for each previously trained model for (K) CPTAC-LUAD, (L) CPTAC-LSCC, (M) CPTAC-PDA, (N) CPTAC-UCEC. Abbreviations: BRCA=breast invasive carcinoma; CRC=colorectal cancer; GBM=glioblastoma; LIHC=liver cancer; LUAD=lung adenocarcinoma; LUSC/LSCC=lung squamous cell carcinoma; OV=ovarian cancer; PAAD/PDA=pancreatic adenocarcinoma; PRAD=prostate adenocarcinoma; UCEC=endometrial cancer

**Supplementary Figure 3:**
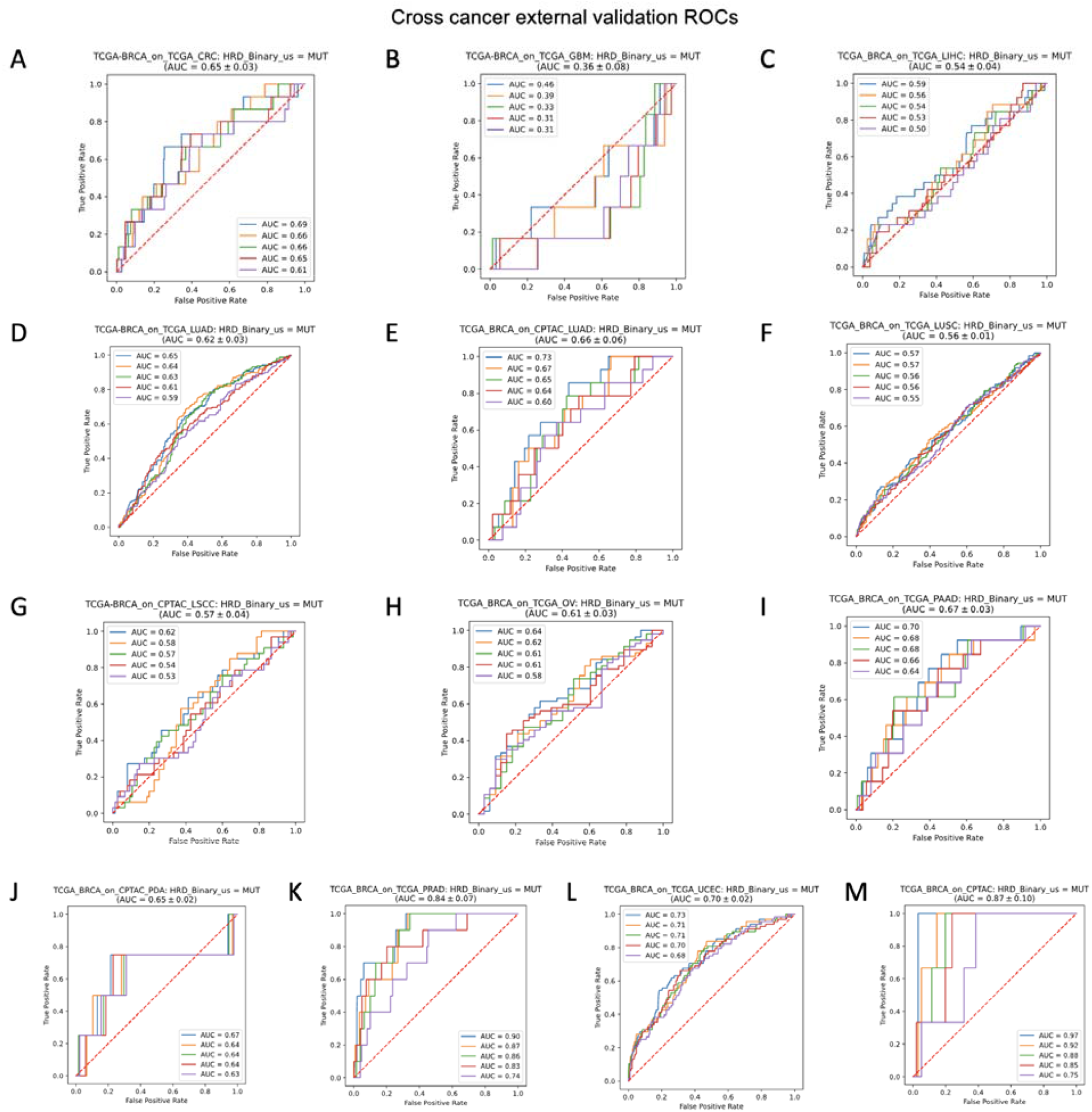
Receiving operating curve for the cross-cancer external validation. The Receiving operating curve (ROC) is shown for the cross-cancer external validation experiment for each model trained on The Cancer Genome Atlas (TCGA) breast cancer (BRCA) cohort for the Homologous recombination deficiency (HRD) binary score on (A) TCGA-CRC, (B) TCGA-GBM, (C) TCGA-LIHC, (D) TCGA-LUAD, (E) CPTAC-LUAD, (F) TCGA-LUSC, (G) CPTAC-LSCC, (H) TCGA-OV, (I) TCGA-PAAD, (J) CPTAC-PDA, (K) TCGA-PRAD, (L) TCGA-UCEC, (M) CPTAC-UCEC. Abbreviations: BRCA=breast invasive carcinoma; CRC=colorectal cancer; GBM=glioblastoma; LIHC=liver cancer; LUAD=lung adenocarcinoma; LUSC/LSCC=lung squamous cell carcinoma; OV=ovarian cancer; PAAD/PDA=pancreatic adenocarcinoma; PRAD=prostate adenocarcinoma; UCEC=endometrial cancer

**Supplementary Figure 4:**
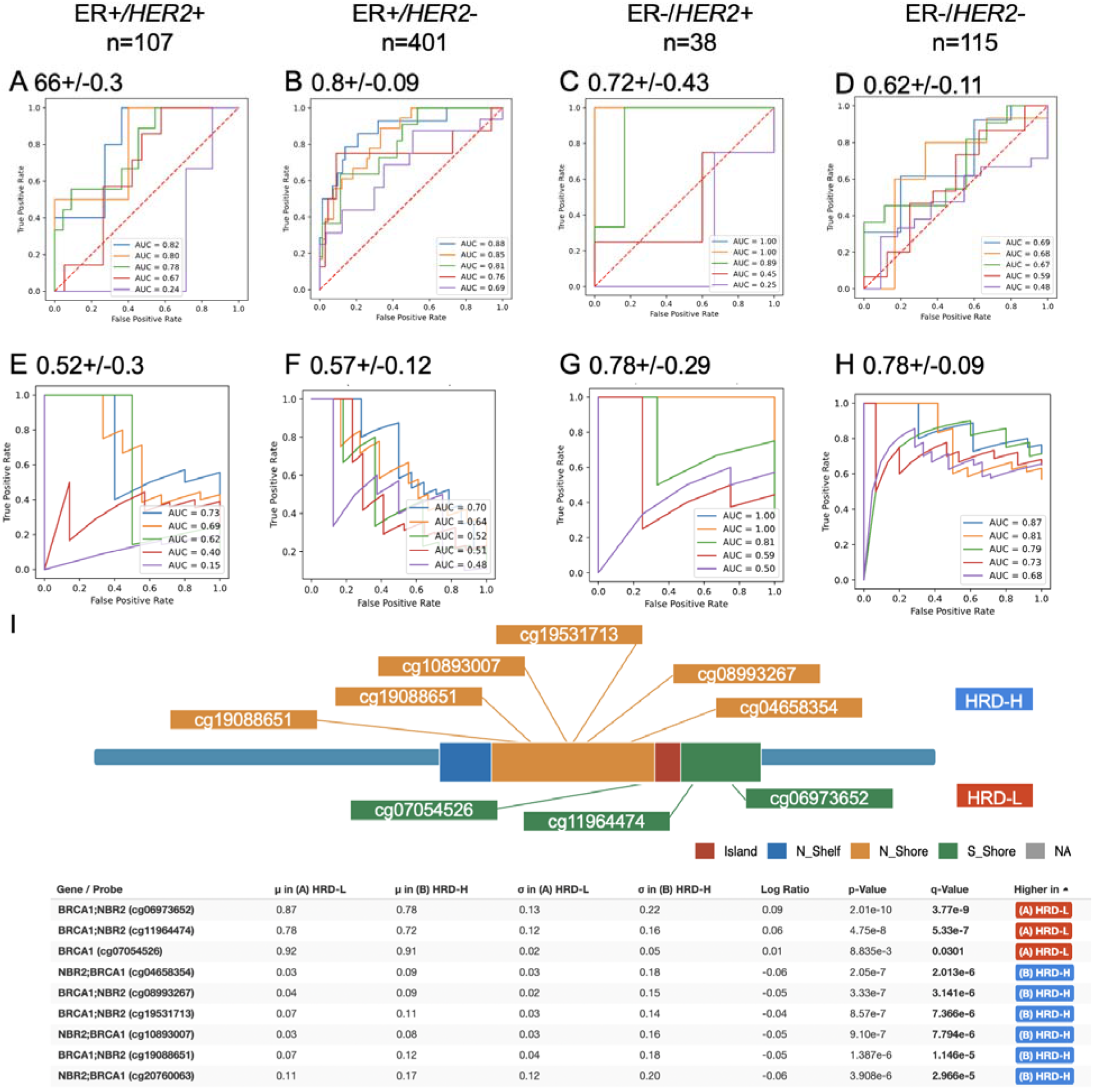
Subgroup analysis and overview the *BRCA1* promotor methylations in TCGA-BRCA. The Receiving operating curve (ROC) and Precision Recall curve (PRC) are shown for the five-fold internal cross-validation experiment for each of the models in The Cancer Genome Atlas - breast cancer (TCGA-BRCA) cohort for the Homologous recombination deficiency (HRD) score. ROC curve is represented for the four different subgroups (A) estrogen receptor positive (ER+) and *HER2*+ ER+ and *HER2*- (C) ER negative (ER-) and *HER2*+ (D) ER- and *HER2*-. The PRC curve is shown for (E) ER+/HER2+, (F) ER+/HER2-, (G) ER-/HER2+, (H) ER-/HER2-. (I) Sketched representation of the occurring promotor methylations (accessed with HM27 and HM450) in the *BRCA1* gene for the ground truth Homologous recombination deficiency high (HRD-H) and low (HRD-L) subgroups.

**Supplementary Figure 5:**
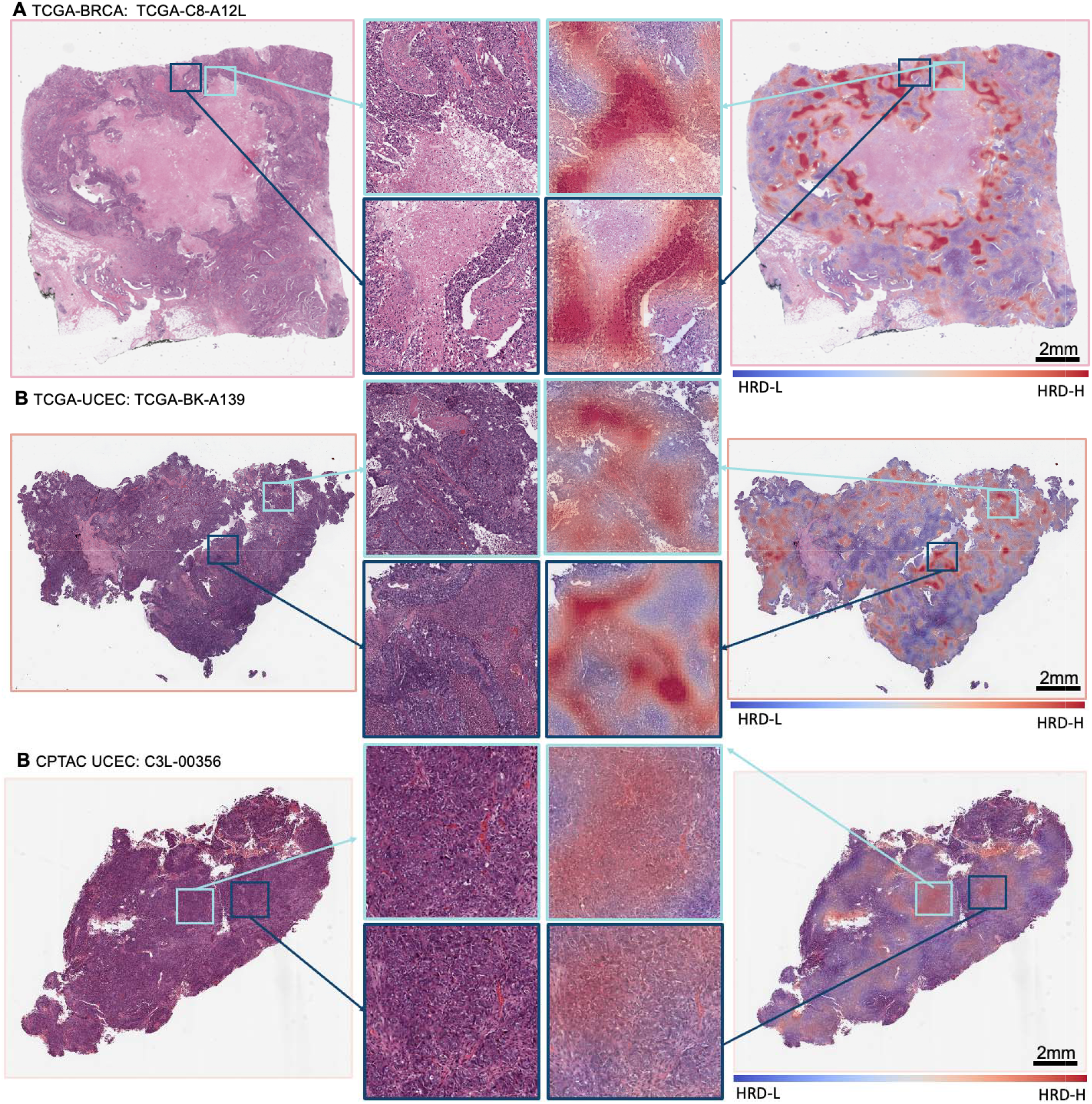
Morphological features of Homologous recombination deficiency in breast and endometrial cancer. Whole Slide Image (WSI) and classification heatmap (ground truth: Homologous recombination deficiency high (HRD-H) and prediction: HRD-H) with magnifications of two different regions. The model was trained on The cancer genome atlas (TCGA) breast cancer (BRCA) cohort and deployed cross cancer wise. Top true positive predicted patients are shown for (A) TCGA-BRCA, (B) Clinical Proteomic Tumor Analysis Consortium (CPTAC) endometrial cancer (UCEC) and TCGA-UCEC.

**Supplementary Figure 6:**
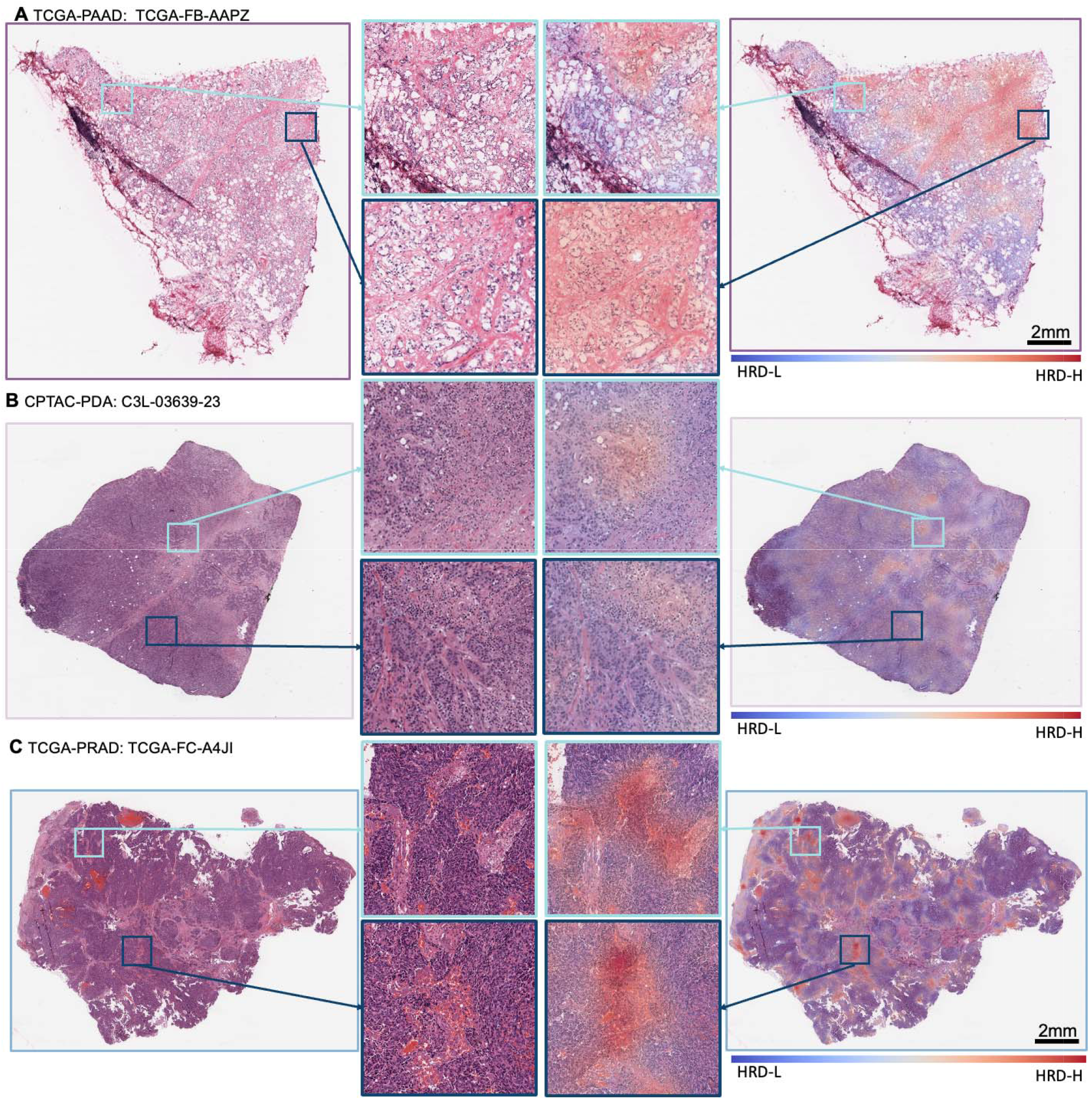
Morphological features of Homologous recombination deficiency in pancreatic and prostate adenocarcinoma. Whole Slide Image (WSI) and classification heatmap (ground truth: Homologous recombination deficiency high (HRD-H) and prediction: HRD-H) with magnifications of two different regions. The model was trained on The cancer genome atlas (TCGA) breast cancer (BRCA) cohort and deployed cross cancer wise. Top true positive predicted patients are shown for (A) TCGA pancreatic adenocarcinoma (PAAD), (B) Clinical Proteomic Tumor Analysis Consortium (CPTAC) pancreatic adenocarcinoma (PDA) and (C) TCGA prostate adenocarcinoma (PRAD).

**Supplementary Table 1: All raw statistical results**. All raw experimental results related to Figure 2, including receiving operating curve (ROC) with 95% confidence interval (CI), Precision-Recall Curve (PRC) with 95% confidence interval (CI), p-values and Homologous recombination deficiency (HRD) high (HRD-H) and HRD-low (HRD-L) patient numbers based on the ground truth, for internal 5-fold cross-validation on The Cancer Genome Atlas (TCGA) external validation on Clinical Proteomic Tumor Analysis Consortium (CPTAC). [Supplementary_Table_1_All_statistical_results.xlsx] in separate file

**Supplementary Table 2: Homologous recombination deficiency score Tables**. Training data and calculated homologous recombination deficiency score (HRD) out of the three subscores loss of heterozygosity (LOH), telomeric allelic imbalance (TAI) and large-scale state transitions (LST) available as continuous (HRDsum) and binary (HRD_Binary) target with a chosen cut off of HRD-L<42 HRD-H>=42 for patients of The Cancer Genome Atlas (TCGA, Sheet1) and Clinical Proteomic Tumor Analysis Consortium (CPTAC, Sheet2).

**Supplementary Table 3: Weblink for customized Homologous recombination deficiency (HRD) subgroups**. Weblink for accessing the clinical and molecular characteristics for both ground truth and prediction Homologous recombination Deficiency (HRD) subgroups at www.cbioportal.org for The Cancer Genome Atlas breast cancer (TCGA-BRCA) Pan Cancer Atlas 2018 study and the TCGA-BRCA Firehose Legacy cohort.

