## Supplementary material for "Direct prediction of Homologous Recombination Deficiency from routine histology in ten different tumor types with attention-based Multiple Instance Learning: a development and validation study": Supplementary_3.docx

Supplementary Table 3

Weblink for accessing the clinical and molecular characteristics for both the customized ground truth and prediction Homologous recombination Deficiency (HRD) subgroups at [www.cbioportal.org](http://www.cbioportal.org) for

1) The Cancer genome Atlas breast cancer (TCGA-BRCA) Pan Cancer Atlas 2018 study

<https://www.cbioportal.org/study/summary?id=brca_tcga_pan_can_atlas_2018#sharedGroups=63e612daabb2dd578e28faf7,63e612e41cec6922c423553a,63eba36d1cec6922c423630e,63eba37c1cec6922c423630f>

2) TCGA-BRCA Firehose Legacy cohort

<https://www.cbioportal.org/study/summary?id=brca_tcga#sharedGroups=63e6129eabb2dd578e28faf5,63e612af1cec6922c4235538>
